# Defining hypoxemia from pulse oximeter measurements of oxygen saturation in well children at low altitude in Bangladesh: an observational study

**DOI:** 10.1101/2021.06.15.21258979

**Authors:** Eric D. McCollum, Carina King, Salahuddin Ahmed, Abu A.M. Hanif, Arunangshu D. Roy, ASMD Ashraful Islam, Tim Colbourn, Holly B. Schuh, Amy Sarah Ginsburg, Shubhada Hooli, Nabidul H. Chowdhury, Syed J.R. Rizvi, Nazma Begum, Abdullah H. Baqui, William Checkley

**Affiliations:** Global Program in Respiratory Sciences, Eudowood Division of Pediatric Respiratory Sciences, Department of Pediatrics, School of Medicine, Johns Hopkins University, Baltimore, Maryland, USA; Health Systems Program, Department of International Health, Johns Hopkins Bloomberg School of Public Health, Baltimore, Maryland, USA; Department of Global Public Health, Karolinska Institutet, Stockholm, Sweden; Projahnmo Research Foundation, Dhaka, Bangladesh; Global Health Institute, University College London, London, United Kingdom; Department of Epidemiology, Johns Hopkins Bloomberg School of Public Health, Baltimore, Maryland, USA; Clinical Trial Center, University of Washington, Seattle, United States; Section of Emergency Medicine, Department of Pediatrics, Baylor College of Medicine, Houston, Texas; Division of Pulmonary and Critical Care, Department of Medicine, School of Medicine, Johns Hopkins University, Baltimore, Maryland, USA; Center for Global Non-Communicable Disease Research and Training, School of Medicine, Johns Hopkins University, Baltimore, Maryland, USA

## Abstract

**Background:** The World Health Organization defines hypoxemia, a low peripheral oxyhemoglobin saturation (SpO_2_), as <90%. Although hypoxemia is an important risk factor for mortality of children with respiratory infections, the optimal SpO_2_ threshold for defining hypoxemia is uncertain in low-income and middle-income countries (LMICs). We derived a SpO_2_ threshold for hypoxemia from well children in Bangladesh residing at low altitude.

**Methods:** We prospectively enrolled well, 3-35 month old children participating in a pneumococcal vaccine evaluation in Sylhet district, Bangladesh between June and August 2017. Trained health workers conducting community surveillance measured the SpO_2_ of children using a Masimo Rad-5® pulse oximeter with a wrap sensor. We used standard summary statistics to evaluate the SpO_2_ distribution, including whether the distribution differed by age or sex. We considered the 2.5^th^, 5^th^, and 10^th^ percentiles of SpO_2_ as possible lower thresholds for hypoxemia.

**Results:** Our primary analytical sample included 1,470 children (mean age 18.6 +/- 9.5 months). Median SpO_2_ was 98% (interquartile range, 96–99%), and the 2.5^th^, 5^th^, and 10^th^ percentile SpO_2_ was 91%, 92%, and 94%. No child had a SpO_2_ <90%. Children 3– 11 months old had a lower median SpO2 (97%) than 12–23 month olds (98%) and 24– 35 month olds (98%) (p=0.039). The SpO_2_ distribution did not differ by sex (p=0.959).

**Conclusion:** A SpO_2_ threshold for hypoxemia derived from the 2.5^th^, 5^th^, or 10^th^ percentile of well children is higher than <90%. If a higher threshold than <90% is adopted into LMIC care algorithms then decision-making using SpO_2_ must also consider the child’s clinical status to minimize misclassification of well children as hypoxemic. Younger children in lower altitude LMICs may require a different threshold for hypoxemia than older children. Evaluating the mortality risk of sick children using higher SpO_2_ thresholds for hypoxemia is a key next step.

**Key Messages:**

- What is the key question? The ideal peripheral oxyhemoglobin saturation (SpO_2_) threshold for defining hypoxemia among children in low-income and middle-income countries is unknown.
- What is the bottom line? A SpO_2_ threshold for hypoxemia set at any of the 2.5^th^, 5^th^, or 10^th^ percentiles of SpO_2_ measurements from well children in a lower altitude setting is higher than the <90% threshold currently recommended by the World Health Organization.
- Why read on? This study is a possible model for other research seeking to establish SpO_2_ thresholds for hypoxemia in children and provides evidence for health policy makers to consider before implementing higher SpO_2_ thresholds than currently in practice in lower altitude settings of low-income and middle-income countries.

## Introduction

Lower respiratory infections (LRIs) kill more young children than any other infectious disease in the world.^1^ The most recent 2017 global estimates report more than 800,000 LRI deaths annually among children below five years of age,^1^ equating to 1-2 deaths every minute. The vast majority of pediatric LRI deaths occur in low-income and middle-income countries (LMICs).^1^ Approximately 30% of all global LRI deaths take place in South Asia each year, and Bangladesh has the 3^rd^ highest annual pediatric LRI incidence and mortality burden among all South Asian countries.^1^

LRIs may be complicated by pulmonary inflammation and areas of ventilation-perfusion mismatch that cause acute hypoxemia, or a low peripheral arterial oxyhemoglobin saturation (SpO_2_) as measured non-invasively by a pulse oximeter.^2^ Acute hypoxemia is an important risk factor for mortality among children with LRIs in LMICs.^3^ For LMICs at lower altitude (i.e., <2,500 meters) the SpO_2_ hypoxemia threshold endorsed by the World Health Organization (WHO) is <90%, a threshold associated with elevated mortality risk among children with LRIs like pneumonia.^3-5^ Per WHO guidelines children with caregiver reported cough and/or difficult breathing accompanied by a SpO_2_ <90% are recommended for hospitalization, parenteral antibiotics, and oxygen administration.^4, 5^ Recent observational studies from Malawi reveal that SpO_2_ thresholds higher than 90% may also be associated with elevated mortality risk among children under five years with clinically diagnosed pneumonia.^6-8^ This evidence suggests the current WHO SpO_2_ hypoxemia threshold of <90% may be suboptimal for identifying higher risk pediatric pneumonia cases for hospitalization in some LMICs.

Despite both the importance and uncertainty around the optimal SpO_2_ threshold for defining hypoxemia few studies from lower altitude settings in LMICs address this issue. One approach commonly used for deriving thresholds for diagnostic tests is to produce a reference range from a healthy population representative of the test’s intended target population.^9, 10^ This approach has been applied to SpO_2_ measurements in children, with most research to date focused on children residing at higher altitudes.^11-15^ In this study we define hypoxemia from the SpO_2_ distribution of well children residing at lower altitude in rural Bangladesh who are participating in a pneumococcal conjugate vaccine (PCV) effectiveness study. We also consider the potential health system implications of implementing a SpO_2_ threshold for hypoxemia derived from a population of well children.

## Methods

### Study design

This is a prospective observational study within a PCV effectiveness evaluation.^16^

### Study setting

Between June and August 2017, the Projahnmo research group, a collaboration between Johns Hopkins University, the Government of Bangladesh’s Ministry of Health and Family Welfare, and Bangladeshi non-governmental and academic institutions, conducted this sub-study in three subdistricts (upazilas) of Syhlet district, northeast Bangladesh. The parent study took place between January 2014 and June 2018.^17^

The Projahnmo research group has a well-established community surveillance system.^17^ The three upazilas under routine surveillance, Zakiganj, Kanaighat, and Beanibazar, are at an altitude between 17 and 23 meters and have a total population of about 770,000 (Figure 1). During routine surveillance local female residents called community health workers (CHWs) visit households within an area of about 10,000 population every two months. At each surveillance visit these trained CHWs provide health counseling to families regarding illness recognition and care seeking, screen women for pregnancy, and evaluate children for respiratory illnesses.

**Figure 1.**
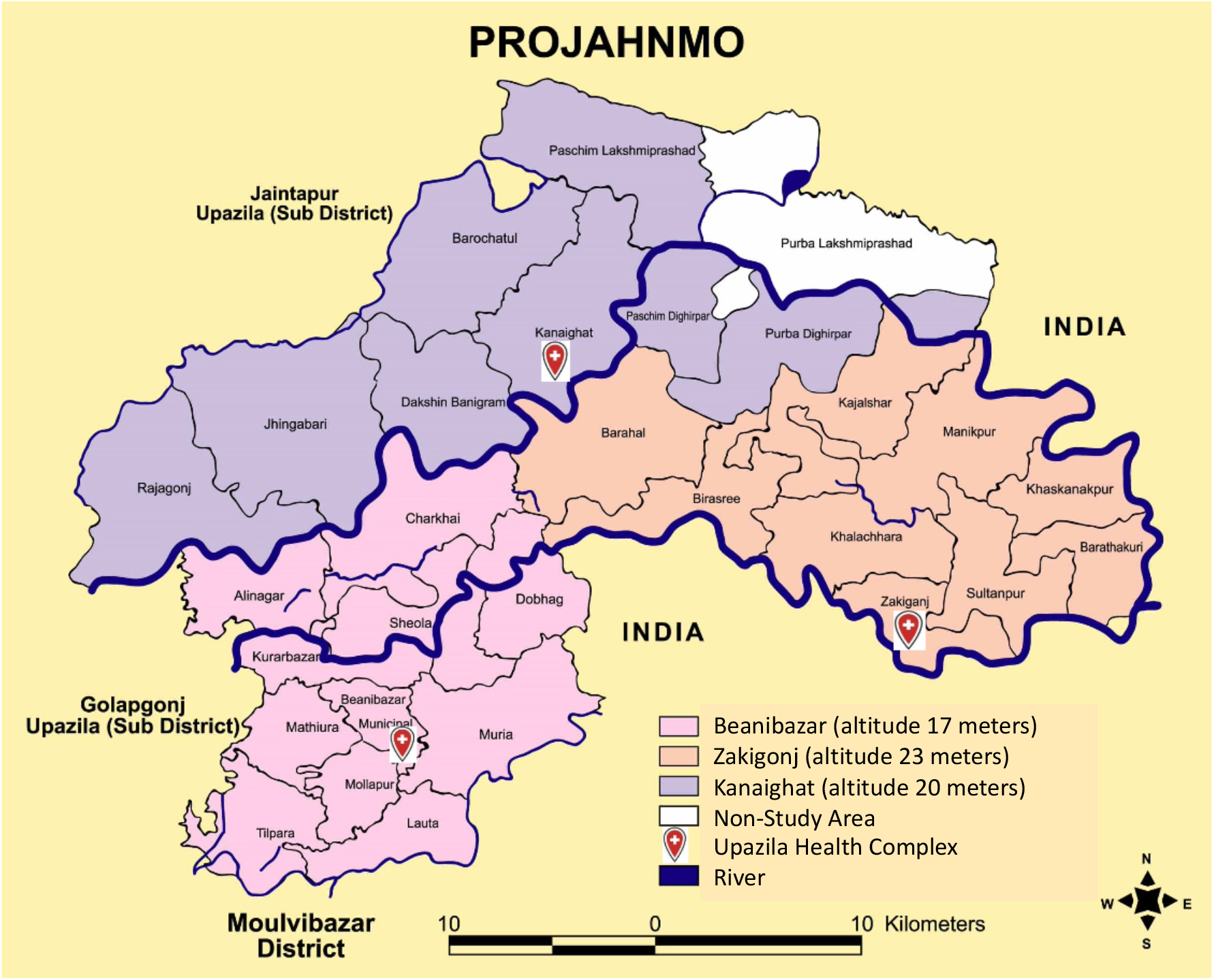
Projahnmo Research Foundation surveillance area.

### Data collection

Twenty-two CHWs were trained in September 2015 to use a Masimo Rad-5® pulse oximeter with a LNCS® Y-I wrap sensor as a part of enhanced respiratory surveillance activities for children during the parent PCV study. The initial training was one day and included theoretical sessions on pulse oximetry supplemented by practice using pulse oximeters to measure the SpO_2_ of volunteer adults and children. During the study period CHWs participated in refresher sessions at least every six months and were routinely supervised by study physicians during household participant screening with the device. Remediation was provided when needed. CHWs were trained to apply the wrap sensor to the big toe of children and gently hold the foot to mitigate movement artifact. SpO_2_ values were considered adequate quality measurements when the CHW achieved the following three metrics; (1) the SpO_2_ value remained stable and non-drifting for no less than three seconds, (2) the quality index signal was of consistent amplitude and displayed at least three green bars, and, (3) the perfusion index signal was at least three green bars in amplitude.

Between June and August 2017 CHWs enrolled well children aged 3-35 months participating in surveillance. CHW screening included an examination for acute signs of an illness and asking caregivers whether the child had any symptoms during the prior week. CHWs observed children for cough, counted the child’s respiratory rate for one minute, measured an axillary temperature with a thermometer, and observed children for any sign of respiratory distress (i.e., head nodding, nasal flaring, audible wheezing, grunting, stridor, tracheal tugging, or lower chest wall indrawing). Children were excluded and referred to the study clinic if aged 3-11 months and had a respiratory rate of ≥50 breaths/minute, or 12-35 months old with a respiratory rate of ≥40 breaths/minute, an axillary temperature >101º F, any vomiting or diarrhea, any WHO-defined general danger sign (lethargy, convulsions, not eating or drinking, severe acute malnutrition), or any sign of respiratory distress as specified above. Children with isolated nasal congestion and/or rhinorrhea were not considered acutely ill and were enrolled.

In order to further filter potentially unwell children from our sample, post-hoc we created three analytic samples from children with a recorded SpO_2_ measurement using different reference heart rate ranges, since an abnormal heart rate may suggest unrecognized illness. Analytic sample 1 is our primary analytic sample, and applies the most conservative estimate of ‘healthy’ with relatively narrow normal heart rate reference ranges of: 120-160 beats/minute for 3-5 month olds, 110-150 beats/minute for 6-11 month olds, 100-140 beats/minute for 12-23 month olds, and 90-130 beats/minute for 24-35 month olds.^18^. Analytic sample 2 is less conservative as it has less restrictive heart rate reference ranges of 100-190 beats/minute for 3-23 month olds and 60-140 beats/minute for 24-35 month olds as normal reference ranges.^18^ Analytic sample 3 ignores heart rate reference ranges altogether and assumes all children are healthy.

### Statistical analysis

Normally distributed continuous variables were described using means and standard deviations, non-normally distributed continuous variables were characterized by medians and interquartile ranges, and bivariate or categorical variables were described using proportions. We considered the 2.5^th^, 5^th^, and 10^th^ percentile of SpO_2_ as possible thresholds for defining hypoxemia. We used the Wilcoxon-Mann-Whitney test for comparisons including a dependent variable without a normal distribution. The Kruskal Wallis test was used for comparisons between a multi-level independent variable and a dependent variable lacking a normal distribution. We fit a linear regression model, adjusted for sex, to explore the association between SpO_2_ and age. Using a power of 80%, significance level of 0.05, and that 25% of children will either be ill, unavailable, or fail measurement, we needed to screen 700 households for each of the three child age strata of 3–11 months, 12–23 months, and 24–35 months (total 2,100) to estimate a mean SpO_2_ of 96% +/- 0.2%. Stata version 16.0 (College Station, TX) was used for all analyses.

### Ethics

The study’s protocol was approved by the Johns Hopkins Bloomberg School of Public Health, Johns Hopkins School of Medicine, Bangladesh Institute of Child Health, and the Ethical Review Committee of the International Centre for Diarrhoeal Diseases Research, Bangladesh, Institutional Review Boards. Written informed consent was obtained from all participant caregivers.

### Patient and public involvement

The development, design, recruitment, conduct, and results of the parent PCV evaluation and this nested study were communicated to the public through local community sensitization meetings held by the Projahnmo study group consortium in Sylhet, Bangladesh.

## Results

### Participant characteristics

From June through August 2017 the CHWs visited 2,098 households and attempted SpO_2_ measurements on 2,042 children (Figure 2). Overall, 20 children with low quality SpO_2_ measurements were excluded at the analysis stage. For primary analytic sample 1 a total of 552 children were additionally omitted due to abnormal heart rates. For analytic sample 2, 157 children were excluded based on reference heart ranges. Among the 1,470 children analyzed for analytic sample 1, the mean age was 18.6 months (SD, 9.5) (Table 1). Average age and the proportion of participants who were female were similar across the three analytic samples.

**Table 1.** Characteristics and SpO_2_ distribution of three analytical samples

|  | Analytic sample 1 (*primary*)* |  |  |  | Analytic sample 2** |  |  |  | Analytic sample 3*** |  |  |  |
| --- | --- | --- | --- | --- | --- | --- | --- | --- | --- | --- | --- | --- |
| Characteristic | All<br>N=1,470 | 3 to 11<br>months<br>N=445 | 12 to<br>23<br>months<br>N=567 | 24 to<br>35<br>months<br>N=458 | All<br>N=1,865 | 3 to 11<br>months<br>N=615 | 12 to<br>23<br>months<br>N=673 | 24 to<br>35<br>months<br>N=577 | All<br>N=2,022 | 3 to 11<br>months<br>N=634 | 12 to<br>23<br>months<br>N=698 | 24 to<br>35<br>months<br>N=690 |
| Age in<br>months,<br>mean (SD) | 18.6<br>(9.5) | 7.3<br>(2.4) | 18.0<br>(3.3) | 30.3<br>(3.2) | 18.2<br>(9.6) | 7.2<br>(2.5) | 18.0<br>(3.3) | 30.2<br>(3.2) | 18.7<br>(9.7) | 7.2<br>(2.5) | 18.0<br>(3.3) | 30.1<br>(3.2) |
| Females, n<br>(%) | 697<br>(47.4%) | 203<br>(45.6%) | 274<br>(48.3%) | 220<br>(48.0%) | 904<br>(48.4%) | 293<br>(47.6%) | 332<br>(49.3%) | 279<br>(48.3%) | 988<br>(48.8%) | 302<br>(47.6%) | 348<br>(49.8%) | 338<br>(48.9%) |
| SpO<sub>2</sub>, 95<sup>th</sup><br>percentile | 100% | 100% | 100% | 100% | 100% | 100% | 100% | 100% | 100% | 100% | 100% | 100% |
| SpO<sub>2</sub>, 75<sup>th</sup><br>percentile | 99% | 99% | 99% | 99% | 99% | 99% | 99% | 99% | 99% | 99% | 99% | 99% |
| SpO<sub>2</sub>, 50<sup>th</sup><br>percentile | 98% | 97% | 98% | 98% | 98% | 97% | 97% | 98% | 98% | 97% | 97% | 98% |
| SpO<sub>2</sub>, 25<sup>th</sup><br>percentile | 96% | 96% | 96% | 96% | 96% | 96% | 96% | 96% | 96% | 96% | 96% | 96% |
| SpO<sub>2</sub>, 10<sup>th</sup><br>percentile | 94% | 94% | 94% | 95% | 94% | 94% | 93% | 94% | 94% | 94% | 93% | 94% |
| SpO<sub>2</sub>, 5<sup>th</sup><br>percentile | 92% | 93% | 92% | 93% | 92% | 92% | 92% | 92% | 92% | 92% | 92% | 92% |
| SpO<sub>2</sub>, 2.5<sup>th</sup><br>percentile | 91% | 92% | 91% | 91% | 91% | 91% | 91% | 91% | 91% | 91% | 91% | 92% |
| SpO<sub>2</sub>, range | 90% to<br>100% | 90% to<br>100% | 90% to<br>100% | 90% to<br>100% | 90% to<br>100% | 90% to<br>100% | 90% to<br>100% | 90% to<br>100% | 90% to<br>100% | 90% to<br>100% | 90% to<br>100% | 90% to<br>100% |

SpO<sub>2</sub> indicates peripheral arterial oxyhemoglobin saturation; SD, standard deviation.
\*Kruskal Wallis test comparing SpO<sub>2</sub> by age category; p value = 0.0387
\*\*Kruskal Wallis test comparing SpO<sub>2</sub> by age category; p value = 0.0095
\*\*\*Kruskal Wallis test comparing SpO<sub>2</sub> by age category; p value = 0.0401

**Figure 2.**
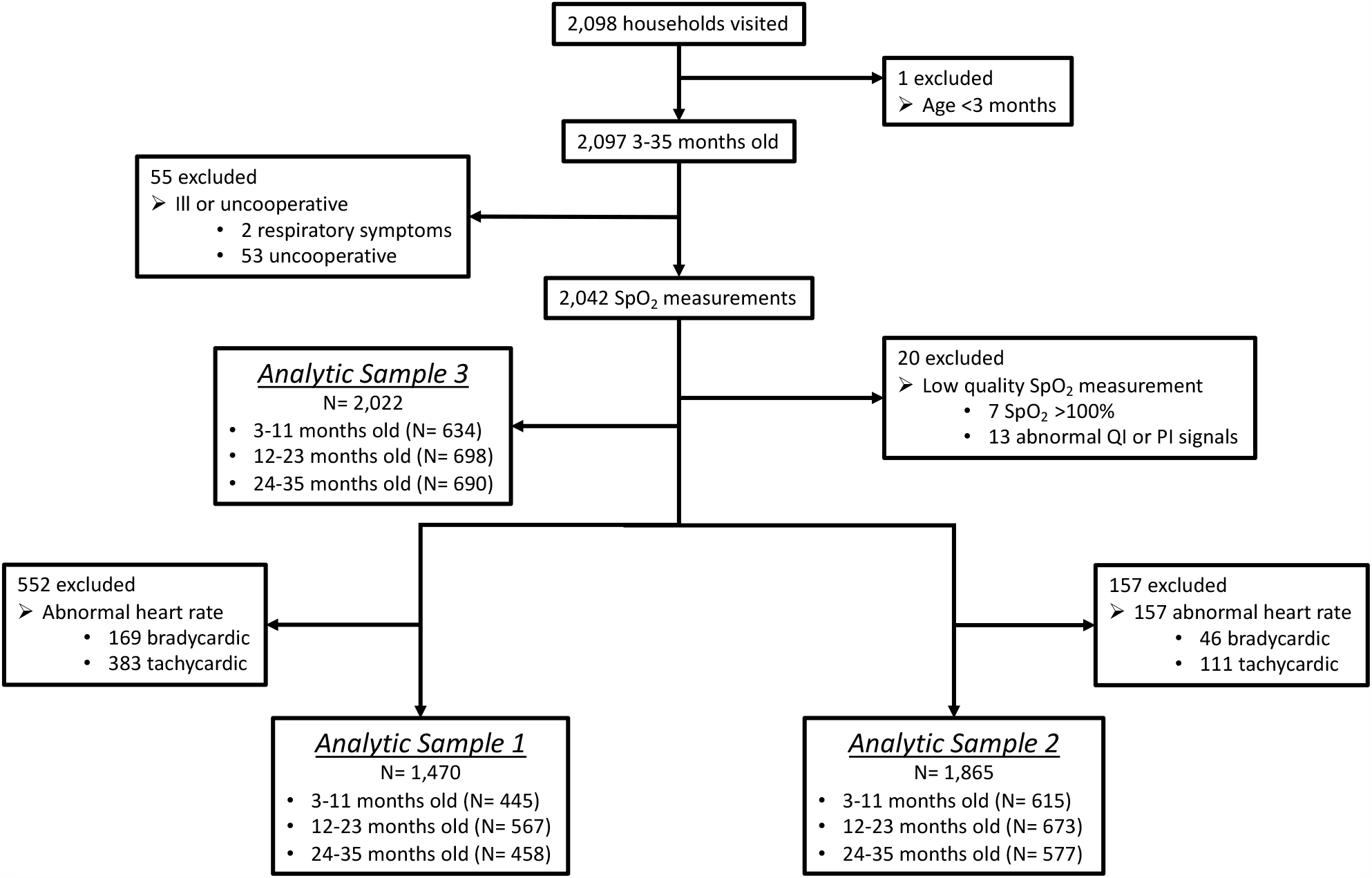
Study profile.

### SpO_2_ distribution

The median SpO_2_ of children included in primary analytic sample 1 was 98% (IQR, 96%, 99%) and the 10^th^, 5^th^, and 2.5^th^ percentile SpO_2_ was 94%, 92%, and 91% (Table 1 and Figure 3). Analytic samples 2 and 3 revealed similar findings (Table 1 and Supplemental Figures 1 and 2). No child included in any of the three analytic samples had a SpO_2_ <90%.

**Figure 3.**
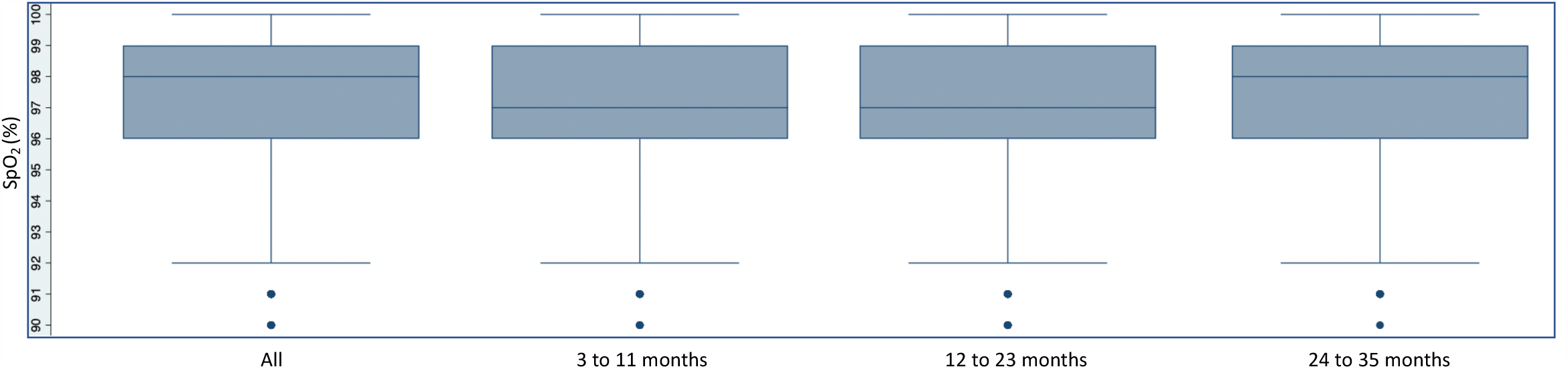
Peripheral oxyhemoglobin saturation (SpO_2_) distribution in Analytic Sample 1.

### Effects of age and sex on SpO_2_

After stratifying measurements into three age strata, 3–11 months, 12–23 months, and 24–35 months, we found children 3–11 months old in primary analytic sample 1 to have a median SpO_2_ of 97%, compared to 98% for each of the two older age strata (p=0.038; Table 1 and Figure 3). We observed similar findings in analytic samples 2 and 3 (Table 1 and Supplemental Figures 1 and 2). When regressing SpO_2_ on age in months, adjusted for sex, we found that for every one month increase in age the SpO_2_ increased by 0.01% (95% CI, 0.001%, 0.02%, p=0.030) in analytic sample 1 (Supplemental Figure 3). We did not observe any difference in the SpO_2_ distribution after stratifying by child sex (p=0.959).

### Health system implications of varying SpO_2_ thresholds

To examine possible consequences on the health system of a SpO_2_ threshold for defining hypoxemia derived from well children we report the probability of a false positive measurement in Table 2 from each analytic sample at differing thresholds. If applying a ≤92% threshold, for example, 76/1,470 (5.1%) well children included in analytic sample 1 would be incorrectly recommended for referral or hospitalization. SpO_2_ thresholds at ≤90%, ≤91%, and ≤93% would incorrectly identify 13 (0.8%), 40 (2.7%), and 117 (7.9%) of the 1,470 children in analytic sample 1 for hospitalization, respectively.

**Table 2.** False positive measurements for hypoxemia by analytical sample and percentile threshold

| Analytic sample | Age in months | 2.5<sup>th</sup> Percentile SpO<sub>2</sub> threshold |  | 5<sup>th</sup> Percentile SpO<sub>2</sub> threshold |  | 10<sup>th</sup> Percentile SpO<sub>2</sub> threshold |  |
| --- | --- | --- | --- | --- | --- | --- | --- |
|  |  | SpO<sub>2</sub> | False positive, n/N (%) | SpO<sub>2</sub> | False positive, n/N (%) | SpO<sub>2</sub> | False positive, n/N (%) |
| 1 | All | 91% | 40/1,470 (2.7%) | 92% | 76/1,470 (5.1%) | 94% | 179/1,470 (12.1%) |
|  | 3-11 | 92% | 20/445 (4.4%) | 93% | 34/445 (7.6%) | 94% | 52/445 (11.6%) |
|  | 12-23 | 91% | 17/567 (2.9%) | 92% | 35/567 (6.1%) | 94% | 83/567 (14.6%) |
|  | 24-35 | 91% | 13/458 (2.8%) | 93% | 29/458 (6.3%) | 95% | 72/458 (15.7%) |
| 2 | All | 91% | 51/1,865 (2.7%) | 92% | 107/1,865 (5.7%) | 94% | 248/1,865 (13.2%) |
|  | 3-11 | 91% | 16/615 (2.6%) | 92% | 33/615 (5.3%) | 94% | 85/615 (13.8%) |
|  | 12-23 | 91% | 21/673 (3.1%) | 92% | 44/673 (6.5%) | 93% | 68/673 (10.1%) |
|  | 24-35 | 91% | 14/577 (2.4%) | 92% | 30/577 (5.1%) | 94% | 60/577 (10.3%) |
| 3 | All | 91% | 55/2,022 (2.7%) | 92% | 119/2,022 (5.8%) | 94% | 273/2,022 (13.5%) |
|  | 3-11 | 91% | 18/634 (2.8%) | 92% | 35/634 (5.5%) | 94% | 88/634 (13.8%) |
|  | 12-23 | 91% | 21/698 (3.0%) | 92% | 48/698 (6.8%) | 93% | 72/698 (10.3%) |
|  | 24-35 | 92% | 36/690 (5.2%) | 92% | 36/690 (5.2%) | 94% | 76/690 (11.0%) |

SpO<sub>2</sub> indicates peripheral arterial oxyhemoglobin saturation.

## Discussion

We derived possible SpO_2_ thresholds for hypoxemia from the 2.5^th^, 5^th^, and 10^th^ percentile cutoffs of well children in rural Bangladesh and estimated the probability of false positive measurements assuming a revised threshold was adopted into care. The SpO_2_ threshold is critical as it triggers a cascade of potentially life or death healthcare decisions and understanding the probability of a false positive SpO_2_ measurement for hypoxemia permits health policy makers to decide how best to balance mortality risk with anticipated hospitalization volumes.

There are two key findings from this research. First, cutoffs for hypoxemia from the 2.5^th^, 5^th^, and 10^th^ percentile are all higher than the current WHO-defined <90% threshold and we did not find any well children below the SpO_2_ <90% threshold. Thus, if any of these cutoffs for hypoxemia are adopted then measuring the SpO_2_ earlier in clinical care pathways when healthier children may be over-represented could increase false positive measurements. This has important implications in health systems with limited resources and potential challenges coping with a higher volume of patient referrals. These results, coupled with findings from Malawi that children with LRI and a SpO_2_ between 90% and 92% are at elevated mortality risk, suggest that the current referral threshold of SpO_2_ <90% minimizes false positives at the expense of false negatives.^6-8^ In order to ensure minimal misclassification of well children as hypoxemic, we recommend care algorithms incorporating a hypoxemia threshold at SpO_2_ levels higher than <90% also consider the child’s clinical status when deciding whether to refer and hospitalize. Second, we found that the SpO_2_ distribution differs by age. Age may therefore need to be considered when establishing a SpO_2_ threshold for hypoxemia. Future analyses that include unwell children should investigate whether this statistical relationship has clinical significance.

To date there have been few attempts to establish a reference range for SpO_2_ measurements of children in LMICs living at lower altitudes. Since most children reside at lower altitudes, understanding the SpO_2_ distribution among this population has broad relevance for identifying the optimal SpO_2_ threshold for hypoxemia. In one study from Chennai, India (altitude 7 meters) the authors measured the SpO_2_ of 626 healthy children aged one month to five years.^19^ In contrast to our findings, the authors found no difference in the SpO_2_ distribution by age. The inclusive 5^th^ percentile of participants in Chennai was a SpO_2_ ≤96%, which was 4% higher than in our study. Other studies that also included healthy children from lower altitudes reported the mean and standard deviation of the SpO_2_ distribution.^13,14,20^ Given the SpO_2_ distribution of healthy children is negatively skewed we described these data using median and percentiles and are therefore unable to make meaningful comparisons.

A more recent multi-country study from the Household Air Pollution Intervention Network (HAPIN) investigators evaluated the SpO_2_ distribution among 1,134 healthy children <24 months old from three lower altitude settings including the same region of India as the prior study (Nagapattinam (altitude 9 meters) and Villupuram (altitude 44 meters), and also in Guatemala (Jalapa District, altitude 1,417 meters) and Rwanda (Kayonza District, altitude 1,354 meters), and one high altitude setting in Puno, Peru (altitude 3,827 meters).^21^ The 5^th^ percentile threshold reported by HAPIN investigators in India was notably consistent with the Chennai study at 96%. Unlike the Chennai study, however, HAPIN investigators found lower 5^th^ percentile thresholds for age in Rwanda (92%) and Guatemala (93%), and observed a correlation between younger age and lower SpO_2_. None of these studies reported the 2.5^th^ percentile cutoff. Overall, it is somewhat surprising that our data from Bangladesh aligns closer with Rwanda and Guatemala than India, another South Asian setting closer in altitude.

Methodology may largely be responsible for the variation in results across these studies. Variation may be due to a combination of device accuracy, including differences in accuracy between devices, variation inherent to measurements on children, measurement variation between healthcare workers and healthcare worker cadres with different training backgrounds, and possible varying degrees of misclassification bias of sick children in each of the three studies. Specifically, the Chennai study used a different pulse oximeter (L&T Medical, Stellar P®) than in the HAPIN study and our study, which both used Masimo devices (Rad-97® and Rad-5®).^19, 21^ Pulse oximeter SpO_2_ estimation algorithms are known to differ by manufacturer and in the United States the Food and Drug Administration requires the testing and certification of pulse oximeters to be accurate within a root mean square error of 3% for arterial blood saturation values between 70% and 100%.^22^ Although data comparing pulse oximeter device performance in children in LMICs is notably limited, it is also well known that device performance can change under conditions common to children like motion and low perfusion.^23,24^

A key methodological difference in this study, compared to the Chennai and HAPIN studies, was formal healthcare workers at healthcare facilities conducting recruitment. In our study trained but informal CHWs recruited and screened children within the community. Although we employed intensive efforts to train and supervise CHWs our use of informal healthcare workers may have influenced both pulse oximeter measurement quality and the number of unwell children remaining in our sample. Our post-hoc data cleaning attempted to further address these possible weaknesses and our findings are reassuringly consistent across the three analytic samples. By contrast, the Chennai and HAPIN studies did not further restrict their analyses by heart rate reference ranges and therefore could remain vulnerable to these issues.

Lastly, although not explicitly stated in either of the Chennai or HAPIN studies, we intentionally did not exclude children with isolated nasal congestion and/or rhinorrhea as anecdotally they are common to otherwise healthy rural Bangladeshi children. In Bangladesh both indoor and ambient air pollution is marked and includes environmental irritants that can cause ongoing upper respiratory mucosal inflammation, nasal congestion, and/or rhinorrhea typical of nonallergic, noninfectious rhinitis.^25^ While it is unlikely that any misclassification bias or device performance inconsistencies were substantially different in our study than in the Chennai and HAPIN studies, it is nevertheless important to interpret our results within this context.

Another issue that may impact the utility of pulse oximeters in all LMICs is the possible greater inaccuracy of SpO_2_ measurements in individuals with darker skin pigmentation.^26^ Although pulse oximetry inaccuracy in individuals with darker skin pigmentation is well established, a recent publication of hospitalized adults in the United States suggests the magnitude and direction of SpO_2_ measurement bias in people with darker skin tones may be clinically unacceptable.^26^ The authors reported a higher odds of hypoxemia – as measured by an arterial blood gas – among darkly pigmented adults with a normal SpO_2_, compared to adults without dark skin pigmentation and a normal SpO_2_.^26^ Further study is needed to better understand the relative contribution and direction of pulse oximeter inaccuracy among children in LMICs where higher percentages of the population often have darker skin pigmentation.

In sum, our findings provide possible reference SpO_2_ thresholds for hypoxemia derived from a population of well children in Bangladesh residing at lower altitude. In fragile, overburdened health systems higher false positive measurements may limit the implementation feasibility of SpO_2_ thresholds above the current <90% mark without additional strengthening of clinical assessments by healthcare providers. This research suggests that age needs to be considered in further work on establishing thresholds for hypoxemia. Key next steps include determining the mortality risk of ill children with SpO_2_ measurements at or below these thresholds in varying LMICs, including Bangladesh, as well as evaluating the performance of SpO_2_ at or below these thresholds for diagnosing LRI. Such research will also shed light on the potential mortality implications of false negative measurements when applying a lower <90% SpO_2_ threshold for hypoxemia.

## Data Availability

Deidentified participant data is available for sharing upon written request and completion of signed data sharing agreements to Dr. Eric D. McCollum at.

## Acknowledgements

We offer our thanks to the caregivers and children participating in this research, as well as to the Projahnmo Study Group field and data management staff, the Ministry of Health and Family Welfare, Government of Bangladesh, GlaxoSmithKline, Bill and Melinda Gates Foundation, and the National Institute of Health their support of this study.

## Contributors

Funding acquisition: AHB and EDM. Conceptualization and design: EDM. Data curation: EDM, AHB, NHC and SJRR. Data collection: EDM, SA, AAMH, ADR, and AI. Data analysis: EDM, NHC, and SJRR. Data interpretation: EDM, CK, SA, AAMH, ADR, AI, TC, HBS, ASG, SH, NHC, SJRR, NB, AHB, and WC. Writing—original draft: EDM. Writing—review & editing: EDM, CK, SA, AAMH, ADR, AI, TC, HBS, ASG, SH, NHC, SJRR, NB, AHB, and WC.

## Funding

This study is funded by the Bill & Melinda Gates Foundation [OPP1084286, OPP1117483] and GlaxoSmithKline [90063241]. EDM was also supported by the Fogarty International Center of the National Institutes of Health under Award Number K01TW009988 for the research reported in this publication. The content is solely the responsibility of the authors and does not necessarily represent the official views of the Bill & Melinda Gates Foundation, GlaxoSmithKline or the National Institutes of Health.

Supplemental Figure 1. Peripheral oxyhemoglobin saturation (SpO_2_) distribution in Analytic Sample 2

Supplemental Figure 2. Peripheral oxyhemoglobin saturation (SpO_2_) distribution in Analytic Sample 3

Supplemental Figure 3. Scatterplot of the relationship between peripheral oxyhemoglobin saturation (SpO_2_) and age in Analytic Sample 1

## Notes

### Competing Interest Statement

The authors have declared no competing interest.

### Clinical Trial

The study did not meet the definition of a clinical trial.

### Author Declarations

The studys protocol was approved by the Johns Hopkins Bloomberg School of Public Health, Johns Hopkins School of Medicine, Bangladesh Institute of Child Health, and the Ethical Review Committee of the International Centre for Diarrhoeal Diseases Research, Bangladesh, Institutional Review Boards. Written informed consent was obtained from all participant caregivers.

