## Supplementary figures and images for "Defining hypoxemia from pulse oximeter measurements of oxygen saturation in well children at low altitude in Bangladesh: an observational study"

### Supplemental Figures 1-3

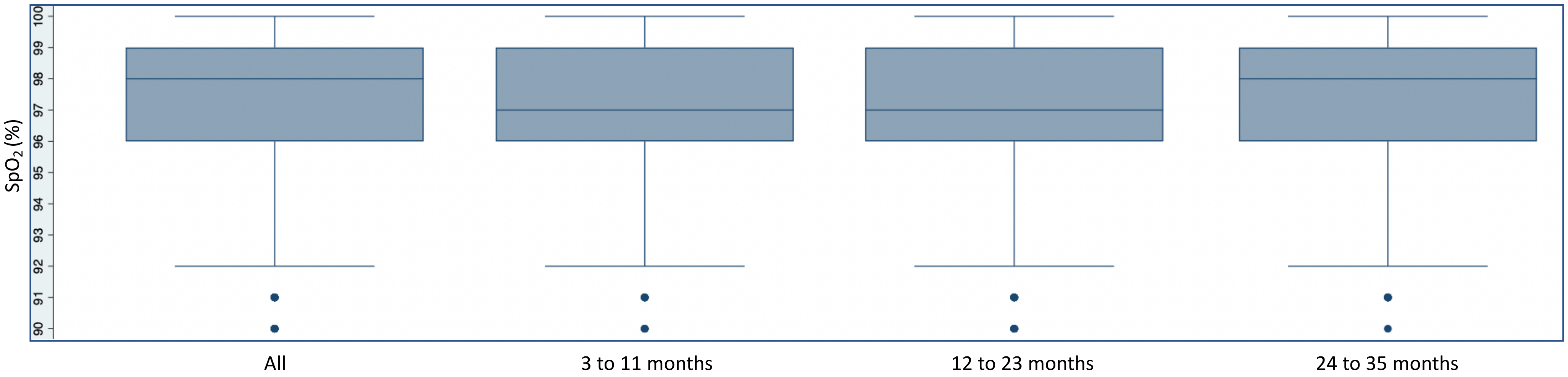

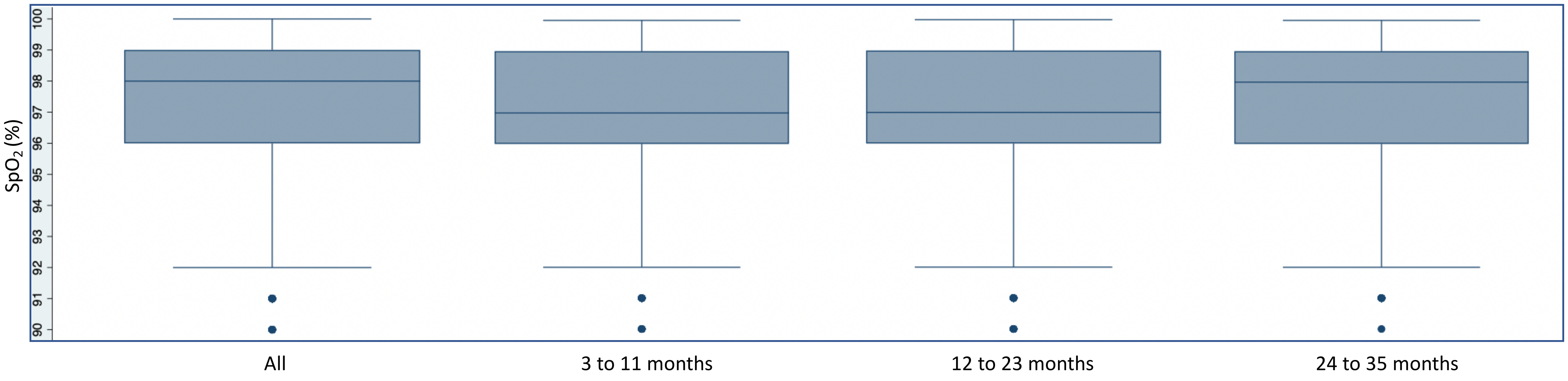

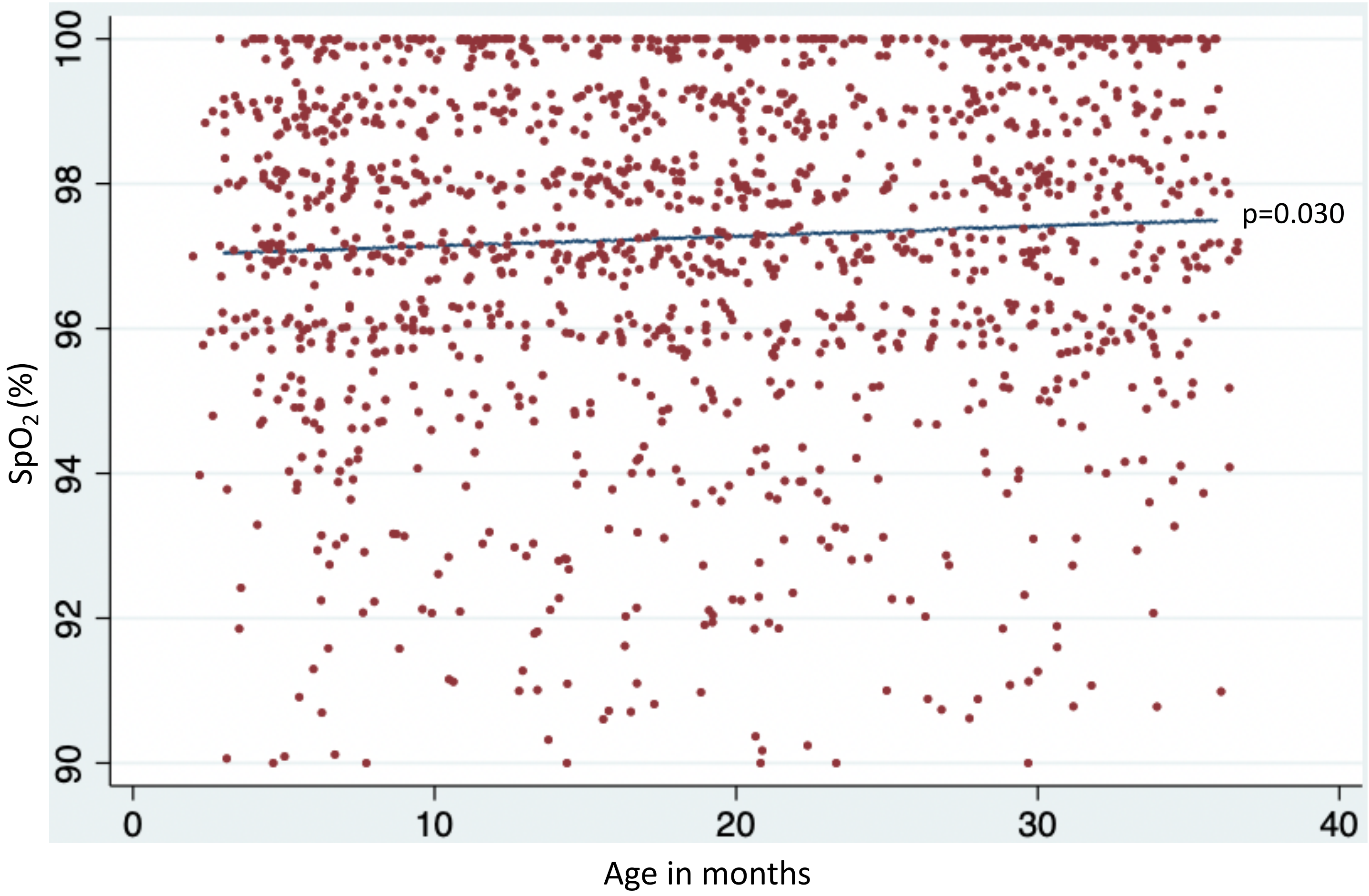
